# Diabetes mellitus, hypertension, and HbA1c, as risk factors for arterial stiffness

**DOI:** 10.1101/2022.01.27.22269098

**Authors:** Rafaela Pelisson Regla, Rogério Toshiro Passos Okawa, Edilson Almeida de Oliveira, Rafael Campos do Nascimento, Milene Cripa Pizatto de Araújo, Giovanna Chiqueto Duarte, Lorena Lima Gargaro, Marina Franciscon Gomes da Cruz, Alex Cardoso Perez, Guilherme Norio Hayakawa, Barbara Letícia da Silva Guedes de Moura, Jorge Juarez Vieira Teixeira

**Affiliations:** Postgraduate Program in Biosciences and Physiopathology, Department of Clinical Analysis and Biomedicine, Health Sciences Center. State University of Maringá, Paraná, Brazil; Department of Medicine, Health Sciences Center. State University of Maringá, Paraná, Brazil

**Keywords:** Hypertension, Diabetes Mellitus, Pulse Wave Velocity, Vascular Stiffness, Risk factors

## Abstract

Studies in a healthy and diabetic population have shown that there is a correlation between the disease and arterial stiffness, and consequently an increase in Pulse Wave Velocity (PWV). We investigate predictors laboratory and clinical associated with arterial stiffness, validated by the increase in PWV. Our findings showed that the predictor for diabetes mellitus, hypertension, glycated hemoglobin ≥5.7, confirmed the significant association for increased arterial stiffness, validated by the increase in PWV.

## INTRODUCTION

Cardiovascular disease (CVD) is the leading cause of death in diabetic patients (Stehouwer *et al*., 2008). Diabetes mellitus (DM) is an independent predictor for cardiovascular adverse events (Farjo *et al*., 2020). Their mechanisms are still not fully understood, but it is believed that increased arterial stiffness is an important mechanism linking diabetes to increased cardiovascular risk (Safar, 1996). Studies have shown that factors related to diabetes and hypertension are associated with a stiffer artery (Motau *et al*., 2018; Resende *et al*., 2019).

A systematic review regarding diabetes, 52% of the studies had a positive association with increased PWV (Cecelja, Chowienczyk, 2009). In another study, the conclusions regarding risk factors and blood pressure were inconsistent or showed low correlation, as these factors would explain only 1% of the PWV variation (Cecelja, Chowienczyk, 2012). In parallel, another research reports that there was no correlation between the variables HbA1c and blood sugar levels in fasting diabetic participants, with arterial stiffness. Concomitantly diabetes and hypertension significantly increased the risk of arterial stiffness (Nuamchit *et al*., 2020).

Currently, great attention is being drawn to the investigation of arterial stiffness, probably due to the evidence that this stiffness, measured by Pulse Wave Velocity (PWV), is an important predictor of cardiovascular events, especially in hypertensive individuals and with other comorbidities (Ben-Shlomo *et al*., 2014; Pierce, 2017). In view of the conflicting results, we investigate predictors laboratory and clinical associated with arterial stiffness, validated by the increase in PWV.

## MTERIAL AND METHODS

All patients of BioCor Cardiology Center, in the city of Maringá, Paraná, Brazil, were subject to a cross-sectional study in the period from 2010 to 2016, using data from a secondary medical records source containing the information of the central pressure of each patient. This research was approved by the Local Ethics Committee (Permanent Ethics Committee in Human Research of the State University of Maringá), approval number 1.664.157/2016.

A non-invasive oscillometric device, the Mobil®-o-graph, was used to measure the central pressure and pulse wave velocity (Jones *et al*., 2000; Luzardo *et al*., 2012; Weiss *et al*., 2012). Three measurements were performed over a 15-minute period, with the subsequent average of these values.

A structured instrument with the independent clinical and laboratory variables: diabetes mellitus – DM (Yes/No), glycated hemoglobin – HbA1c (<5.7, ≥5.7–6.4, ≥6.5%)^a,b^, systemic arterial hypertension – SAH (Yes/No), total cholesterol (<190, ≥190 mg/dl), low-density lipoproteins - LDLc (< 130, ≥ 130 mg/dl), high-density lipoproteins – HDLc (<40, ≥40 mg/dl), triglycerides (<150, ≥150 mg/dl) was used to collect data. The PWV was considered as an outcome variable with two cuts, <10 and ≥10 m/s.

The analyses were conducted using Stata 9.0 (StataCorp, College Station, TX 77845 USA). The statistical analysis for variables was validated with p-value <0.05 by the chi-square test, and an Odds Ratio (OR) measure and a 95% confidence interval were used. In the logistic model construction for the primary outcome of PWV, all variables regardless of the p-value were considered in the analysis and adjusted for DM, hypertension, HbA1c, total cholesterol, LDLc, HDLc d LDL and Triglycerides.

## RESULTS

The population considered in this study was 1197 patients, and 341 (28.5%) of these patients had altered PWV and 856 (71.5%) not had altered PWV. Table I shows the univariate analysis of the risk factors for pulse wave velocity between the two PWV groups <10 and ≥10, using association effect measures, estimated by the Odds Ratio. The variables impacting for PWV ≥ 10, with statistical significance for p <0.001 were: DM, hypertension, HbA1c ≥ 5.7, total cholesterol ≥ 190 mg/dl, LDL cholesterol ≥ 130 mg / dl, and HDL cholesterol ≤ 40 md/dl.

**TABLE I.**
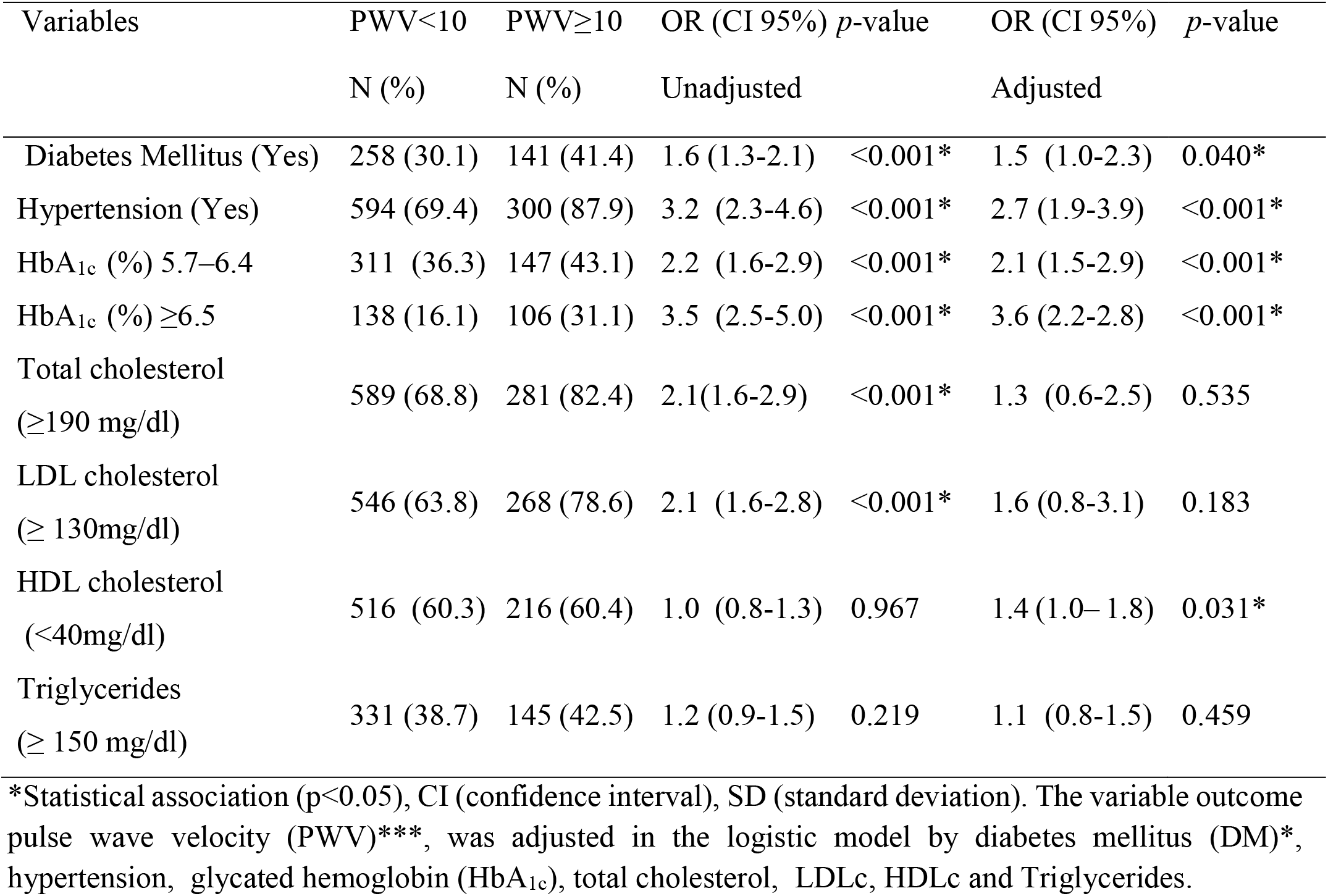
Logistic regression of clinical and laboratory risk factors for arterial stiffness

Next, a logistic regression analysis tried to verify the association of independent variables adjusted for <0.05 and the outcome variable PWV. The final model showed a positive association for PWV ≥10 for the presence of DM (OR 1.5, CI 1.0-2.3, p=0.040), hypertension (OR 2.7, CI 1.9-3.9, p <0.001), HbA1c 5.7-6.4 (OR 2.1, 1.5-2.9, p<0.001), HbA1c ≥ 6.5 (OR 3.6, 2.2-2.8, p<0.001), and HDL cholesterol ≤ 40 md/dl (OR 1.4, IC 1.0-1.8, p=0.031).

## DISCUSSION

The study’s findings showed that diabetes mellitus, arterial hypertension and elevated glycated hemoglobin are predictors for arterial stiffness, validated by the PWV. Our study values ≥ that 5.7% of HbA1c showed an estimated risk gradient increasing between 110 to 260% for PWV alteration than patients with values lower than 5.7%. Studies show a strong relationship between high levels of glycated hemoglobin and an increased risk of cardiovascular diseases (Kilpatrick, Bloomgarden, 2013; Mannucci *et al*., 2009). This result could also be observed in another study where elevated glycated hemoglobin was positively associated with elevated PWV, that is, lousy diabetes control would lead to greater arterial stiffness (Chen *et al*., 2009). Although xx has not shown a correlation between arterial stiffness and HbA1c and fasting blood sugar levels in diabetic participants, the authors noted that concomitant diabetes and hypertension significantly increased the risk of arterial stiffness (Nuamchit *et al*., 2020).

In individuals with primary HA, the risk for any cardiovascular (CV) complication increases in parallel to the elevation of the PWV (DeLoach & Townsend, 2008), as vessel stiffening is accelerated in the presence of the disease(Kotsis *et al*., 2011). The results obtained demonstrate that hypertension patients are approximately two times more likely to increase PWV than patients with no hypertension. The most important modification that occurs in the vessel wall of a hypertensive patient is the hypertrophy of the middle layer associated with reduced arterial compliance and distensibility regardless of the BP level (Benetos *et al*., 2002; Blacher *et al*., 1999).

The study has some limitations, but also good points. The first limitation refers to the size of the population involved. Despite the robust sample of 1197 patients, larger samples would provide greater precision for the evidence’s findings and conclusion, specifically about to with concerning to the diabetes mellitus predictor. The second limitation concerns the research’s performance in one single ambulatory service, considering the difficulty or lack of technique for PWV assessment. The sample period is also a point to be valued since the analysis and research with these patients took six years. Despite the limitations or possible biases presented in the study, we believe that the findings can be relevant to the medical clinic and health services.

## CONCLUSION

The predictor’s diabetes mellitus, hypertension, glycated hemoglobin ≥5.7 adjusted by logistic regression analysis, confirmed a significant association with increased arterial stiffness, validated by the increase of pulse wave velocity.

## Data Availability

All data produced in the present work are contained in the manuscript

